# Broad neutralization of SARS-CoV-2 variants, including omicron, following breakthrough infection with delta in COVID-19 vaccinated individuals

**DOI:** 10.1101/2021.12.01.21266982

**Authors:** Thomas Lechmere, Luke B. Snell, Carl Graham, Jeffrey Seow, Zayed A. Shalim, Themoula Charalampous, Adela Alcolea-Medina, Rahul Batra, Gaia Nebbia, Jonathan D. Edgeworth, Michael H. Malim, Katie J. Doores

**Author notes:** These authors contributed equally (Thomas Lechmere, Luke B. Snell).

## Abstract

Numerous studies have shown that a prior SARS-CoV-2 infection can greatly enhance the antibody response to COVID-19 vaccination, with this so called “hybrid immunity” leading to greater neutralization breadth against SARS-CoV-2 variants of concern. However, little is known about how breakthrough infection (BTI) in COVID-19 vaccinated individuals will impact the magnitude and breadth of the neutralizing antibody response. Here, we compared neutralizing antibody responses between unvaccinated and COVID-19 double vaccinated individuals (including both AZD1222 and BNT162b2 vaccinees) who have been infected with the delta (B.1.617.2) variant. Rapid production of Spike-reactive IgG was observed in the vaccinated group providing evidence of effective vaccine priming. Overall, potent cross-neutralizing activity against current SARS-CoV-2 variants of concern was observed in the BTI group compared to the infection group, including neutralization of the omicron (B.1.1.529) variant. This study provides important insights into population immunity where transmission levels remain high and in the context of new or emerging variants of concern.

## Introduction

COVID-19 vaccines have proven to be critical in controlling SARS-CoV-2 infections worldwide. Vaccines based on the SARS-CoV-2 Wuhan-1 Spike protein generate neutralizing antibodies which constitute an important component of the protective capacity of COVID-19 vaccines. Since the beginning of the global pandemic, variants of SARS-CoV-2 have arisen which encode mutations in the Spike protein. Until November 2021, the dominant circulating variant was B.1.617.2 (delta), but B.1.1.529 (omicron) is rapidly increasing globally (https://www.nicd.ac.za/wpcontent/uploads/2021/11/Update-of-SA-sequencing-data-from-GISAID-26-Nov_Final.pdf). There is concern that SARS-CoV-2 variants of concern (VOCs) might lead to a reduction in vaccine efficacy, in particular against omicron which encodes 31 amino acid changes in the Spike protein.

To generate high titres of Spike reactive IgG with potent neutralization, double vaccination is required for both the BNT162b2 (based on mRNA encoding a stabilized Spike) and AZD1222 (based on a chimp adenovirus encoded Spike) vaccines (Ramasamy et al., 2021; Walsh et al., 2020). Importantly, several studies have shown that SARS-CoV-2 infection prior to vaccination can boost antibody titres and neutralizing activity, with this so called “hybrid immunity” leading to greater neutralization breadth against SARS-CoV-2 VOCs (Goel et al., 2021; Manisty et al., 2021; Reynolds et al., 2021; Saadat et al., 2021; Stamatatos et al., 2021). However, little is known about how breakthrough infection (BTI) in COVID-19 double vaccinated individuals will impact the magnitude and breadth of the neutralizing antibody response (Collier et al., 2021; Hacisuleyman et al., 2021; Kitchin, 2021), particularly in the face of the omicron variant where preliminary data shows that a 3^rd^ vaccine dose is required for robust neutralization activity (Cameroni, 2021; Doria-Rose, 2021; Garcia-Beltran, 2021; Gruell, 2021; Schmidt, 2021) and predicted for high vaccine efficacy (Khoury, 2021). This information would provide important insights into population immunity in areas where transmission levels remain high and where omicron is rapidly becoming the dominant strain. Here, we compared the magnitude and breadth of the antibody response in individuals infected with the SARS-CoV-2 delta VOC (vaccine naïve) to the antibody response in individuals who were double vaccinated prior to delta infection (breakthrough infection, BTI).

## Results

### Cohort description

We identified 42 individuals admitted to St Thomas’ hospital who had previously received two COVID-19 vaccinations and subsequently tested positive for COVID-19. We note that at the time of writing, from the patients admitted to St Thomas’ Hospital with COVID-19 since the emergence of delta (n=635), 260 cases out of 332 (78%) where vaccination was known were either unvaccinated or partially vaccinated (one inoculation). In this study, 30/42 (71%) of patients in the BTI group were admitted to hospital due to COVID-19, of which 11/30 (37%) patients experienced severe disease (severity 4-5). 29/30 (97%) patients had underlying health conditions that predispose to severe disease and aged between 20-103 years (median age 77 years, IQR 59-86) (**Table S1**). The remaining 12 participants in the BTI group (29%) were asymptomatic and admitted for reasons other than COVID-19. Patients were aged between 24-96 years (median age 62 years, IQR 37-72). Overall, the BTI group included individuals receiving both the AZD1222 vaccine (n = 23) and the BNT162b2 vaccine (n = 19). Discarded serum samples were collected between 0-53 days post onset of symptoms (POS) and longitudinal serum samples were collected where possible. The number of days post second vaccine ranged from 29-179 days (median 109 days).

Sera (n = 19) were also collected from unvaccinated individuals admitted to St Thomas’ hospital due to COVID-19 who had a confirmed infection with the SARS-CoV-2 delta variant and experienced a range of disease severities with 9/19 (47%) patients experiencing severe disease (severity 4-5). Patients were aged between 25-82 years (median age 39 years, IQR 30-51) and 9/19 (47%) had underlying health conditions (**Table S2**). Sera were collected between 12-22 days POS.

### IgG and IgM to Spike in breakthrough infection

First, we measured the IgG and IgM responses to recombinant Spike (both WT and delta) in the two groups by ELISA. Sera from unvaccinated individuals infected with the delta variant at 12-22 days POS had higher delta Spike IgM levels than delta Spike IgG (**Figure 1A**) indicative of a primary immune response. Slightly higher IgG and IgM titres were observed against the delta recombinant Spike compared to WT Spike (**Figure 1B**).

**Figure 1:**
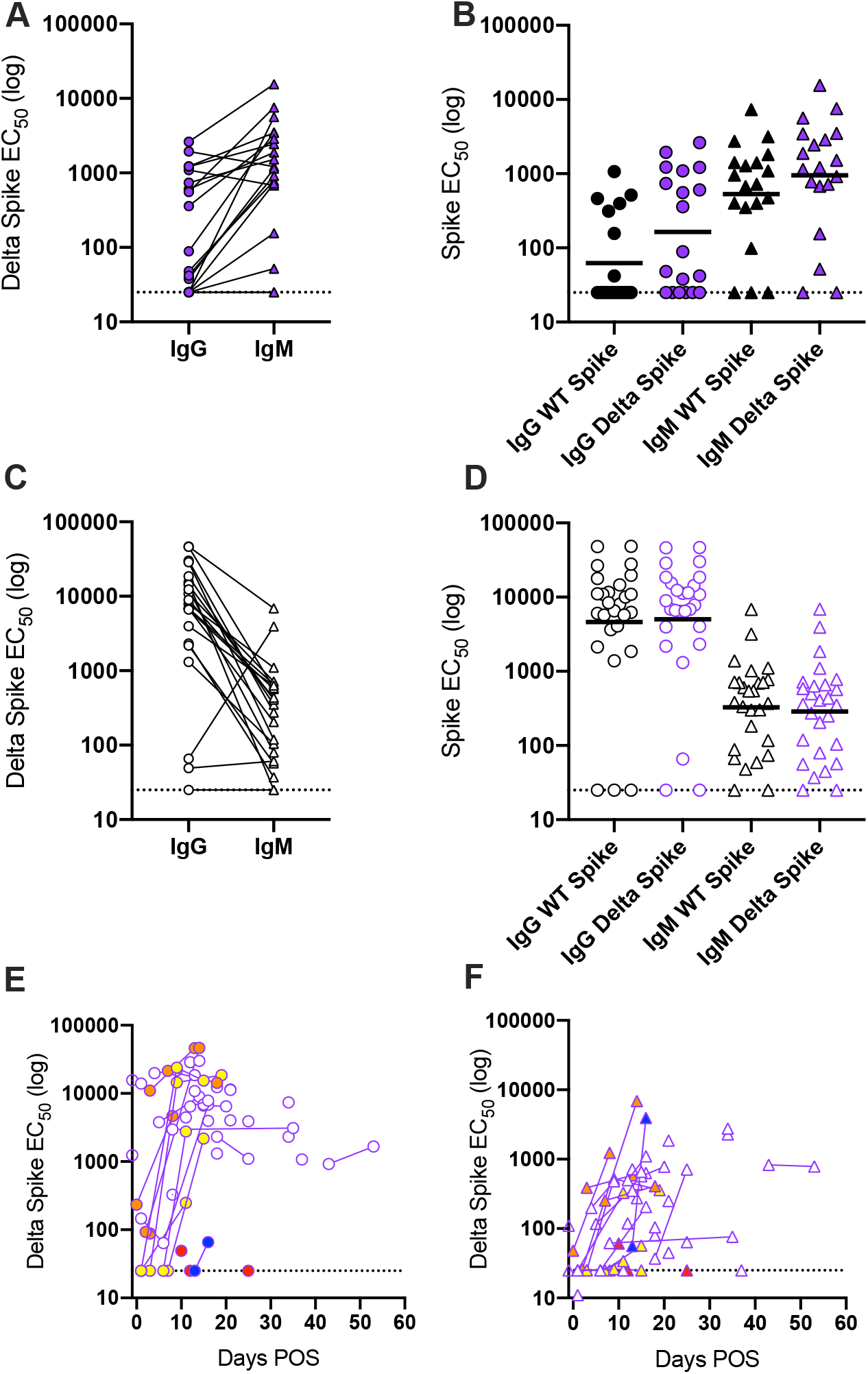
Differences in antibody binding between delta infected individuals and COVID-19 vaccinated individuals experiencing delta breakthrough infection. **A)** Difference in IgG and IgM titres for sera collected 12-22 days POS for the delta infection (vaccine naïve) group. **B)** Comparison of the IgG and IgM ED_50_ values against recombinant WT and delta Spikes for the vaccine naïve group. Black horizontal lines show the geometric mean titres. **C)** Difference in IgG and IgM titres for sera for the BTI group. **D)** Comparison of the IgG and IgM ED_50_ values against recombinant WT and delta Spikes for the BTI group. Black horizontal lines show the geometric mean titres. **E)** Longitudinal IgG ED_50_ against recombinant delta Spike in the BTI group. **F)** Longitudinal IgM ED_50_ against recombinant delta Spike in the BTI group. Donors with IgM>IgG are shown in blue, donors who do not seroconvert are shown in red, donors with high Spike IgG but no N IgG at <7days POS are shown in orange, and donors with low Spike IgG at <7 days POS that rapidly increases are shown in yellow.

For sera collected 12-22 days POS in the BTI group, delta Spike IgM levels in the BTI group were lower than the delta Spike IgG level (**Figure 1C**) indicative of a recall response. A similar trend was observed for both AZD1222 and BNT162b2 vaccinated individuals (**Figure S1A**). Where sequential serum samples were collected, nine individuals had undetectable or a very low Spike IgG response at the earliest timepoint POS (**Figure 1D and Figure S1B**). However, high titres of Spike specific IgG were detected several days later with only modest increases in IgM titres (**Figure 1E-F**). Six donors had IgG against Spike at early time points but lacked IgG to the SARS-CoV-2 Nucleoprotein (**Figure S1C-D**). Although this may provide insight into Spike IgG levels prior to infection, it is more likely due to a rapid Spike IgG recall response compared to a de novo IgG response to Nucleoprotein (**Figure 1E-F)**. One participant (a renal transplant patient) had a high IgM response and low IgG response, similar to the vaccine naïve group, which suggests failed vaccine priming (**Figure 1C**). Interestingly, unlike the vaccine naïve group, the IgG and IgM titres against the WT and delta Spikes were comparable in the BTI group (**Figures 1D**).

Overall, these results indicate a rapid recall response due to prior vaccination in the BTI group and a primary immune response in the vaccine naïve group.

### Neutralization activity following breakthrough infection

Next, we measured neutralization breadth and potency in the two groups using HIV-1 (human immunodeficiency virus type-1) based virus particles, pseudotyped with SARS-CoV-2 Spike from different VOCs (wild-type (Wuhan), alpha (B.1.1.7), delta (B.1.617.2), mu (B.1.621) and beta (B.1.351)) and a HeLa cell-line stably expressing the ACE2 receptor (Seow et al., 2020). The majority (17/19, 89%) of the vaccine naïve group produced a robust homologous neutralizing response against the delta VOC (**Figure 2A**). Cross-neutralization of the parental strain and other VOCs was detected for most individuals, albeit at a reduced potency. As we have reported previously (Dupont et al., 2021), the greatest reduction was observed against beta with a 9.4-fold reduction in the GMT, reflecting greater antigenic distance.

**Figure 2:**
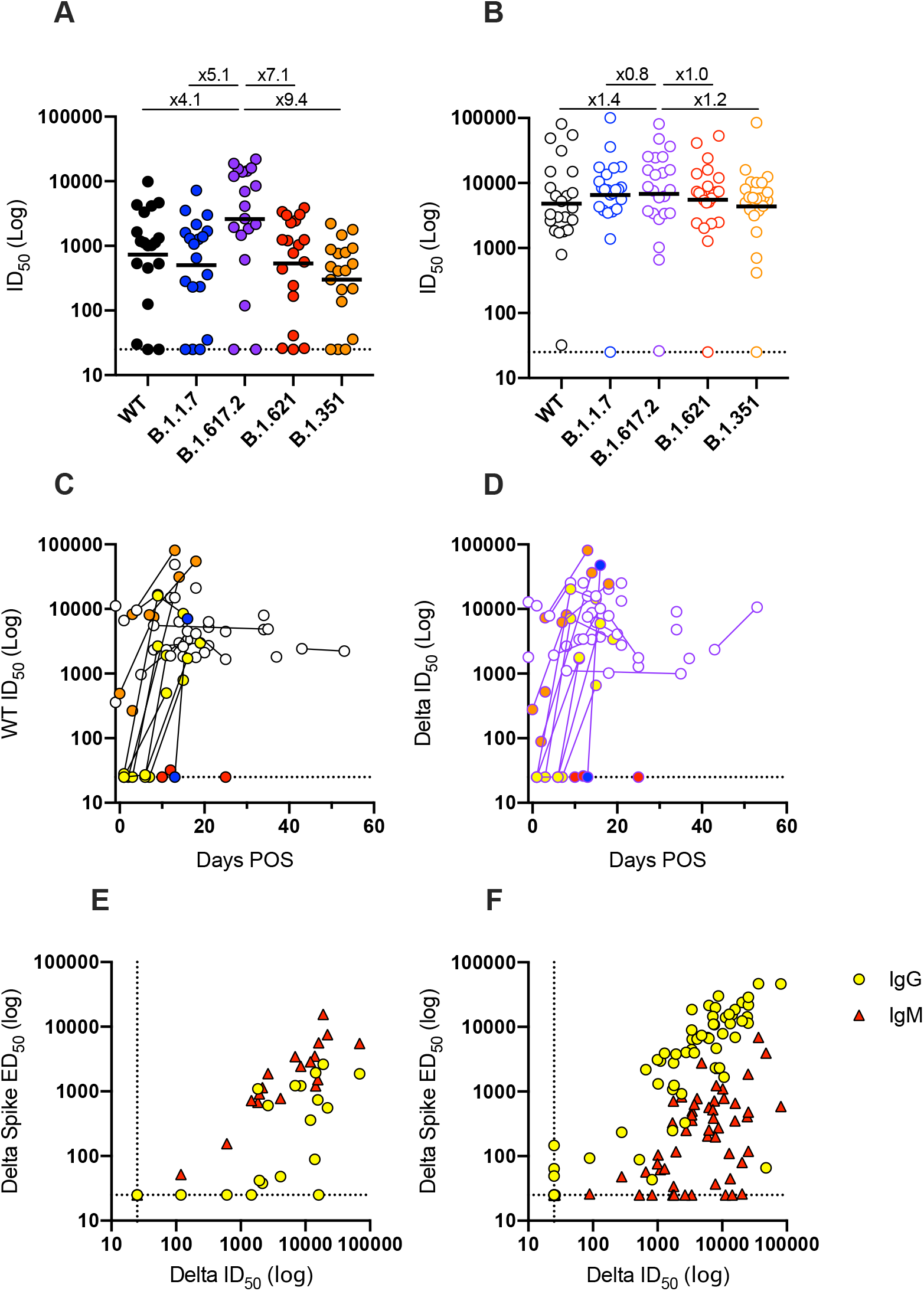
Differences in neutralizing antibody response between delta infected individuals and COVID-19 vaccinated individuals experiencing delta breakthrough infection. ID_50_ of neutralization against WT (black) and VOCs alpha (blue), delta (purple), mu (red) and beta (orange) for sera from **A)** SARS-CoV-2 delta infected individuals and **B)** BTI individuals. Black line shows the geometric mean titre. Fold decrease in GMT compared to delta are shown above. Longitudinal neutralization potency of sera in BTI individuals against **C)** WT pseudovirus particles and **D)** delta pseudovirus particles. Donors with IgM>IgG are shown in blue, donors who do not seroconvert are shown in red, donors with high Spike IgG but no N IgG at <7days POS are shown in orange, and donors with low Spike IgG at <7 days POS that rapidly increases are shown in yellow. Data for the alpha, beta and mu VOCs is shown in **Figure S2A**. Correlation (Spearman, r) between ID_50_ of neutralization and IgM or IgG ED_50_ for delta Spike binding for **E)** delta infected individuals (IgM: r = 0.92, r^2^ = 0.90, p <0.0001 and IgG: r = 0.66, r^2^ = 0.43, p = 0.001) and **F)** COVID-19 vaccinated individuals experiencing breakthrough infection (IgM: r = 0.61, r^2^ = 0.38, p <0.0001 and IgG: r = 0.83, r^2^ = 0.75, p <0.0001). A linear regression was used to calculate the goodness of fit (r^2^). The dotted lines represent the lowest serum dilution used in each assay. IgG is shown with yellow circles and IgM shown with red circles.

Sera collected between 12-22 days POS from individuals in the BTI group showed a robust homologous neutralizing response as well as strong cross-neutralization of the parental variant and VOCs (**Figure 2B**). Only a 1.2-fold reduction in GMT was observed against the more neutralization resistant beta VOC. Several individuals in the BTI group with sera collected soon after onset of symptoms showed no or very low neutralization against both WT and delta variants, however, potent neutralizing activity was detected several days later (**Figure 2C&D and S2A**). Geometric mean titres against the five variants were very similar between AZD1222 and BNT162b2 vaccinated individuals (**Figure S2B**). Three participants in the BTI group either failed to produce neutralizing antibodies or had titres close to baseline despite vaccination and SARS-CoV-2 infection. These individuals had underlying health conditions including cancer (one participant was undergoing rituximab treatment) and type-2 diabetes.

As would have been anticipated, IgG ED_50_ values correlated best with ID_50_ values for the BTI group whereas IgM ED_50_ values correlated best with ID_50_ values for the unvaccinated group (**Figure 2E&F**) further highlighting the priming capacity of both the AZ and BNT162b2 vaccines.

### BTI generates neutralizing activity against omicron

In November 2021, omicron (B.1.1.529) was identified that encoded 31 amino acid mutations in the Spike protein (**Figure 3A**). Initial studies suggest that these mutations lead to large reductions in neutralization of sera from double vaccinated individuals. However, administration of a third vaccine dose greatly enhances neutralization titres against omicron suggesting incomplete neutralization escape (Cameroni, 2021; Cele et al., 2021; Doria-Rose, 2021; Garcia-Beltran, 2021; Gruell, 2021; Schmidt, 2021). Neutralization activity of a subset of 14 sera from the vaccine naïve group and 15 sera from the BTI group were measured against WT, delta and omicron variants (**Figure 3B&D**). In delta infected individuals, a 28.9-fold reduction in GMT against omicron compared to delta was measured compared to a 6.9-fold reduction in GMT for WT. Sera from two participants did not neutralize the omicron variant at the lowest dilution point (1:25). In contrast, all 15 sera from the BTI group neutralized the omicron variant with only a 4.5-fold reduction in GMT against omicron compared to delta GMT (**Figure 3C&E**). Three individuals showed a 21-to 81-fold reduction in ID_50_ against omicron compared to ID_50_ against delta, all of which were receiving treatment for underlying health conditions. These results further highlight the breadth of the neutralizing antibody response following BTI with the delta variant.

**Figure 3:**
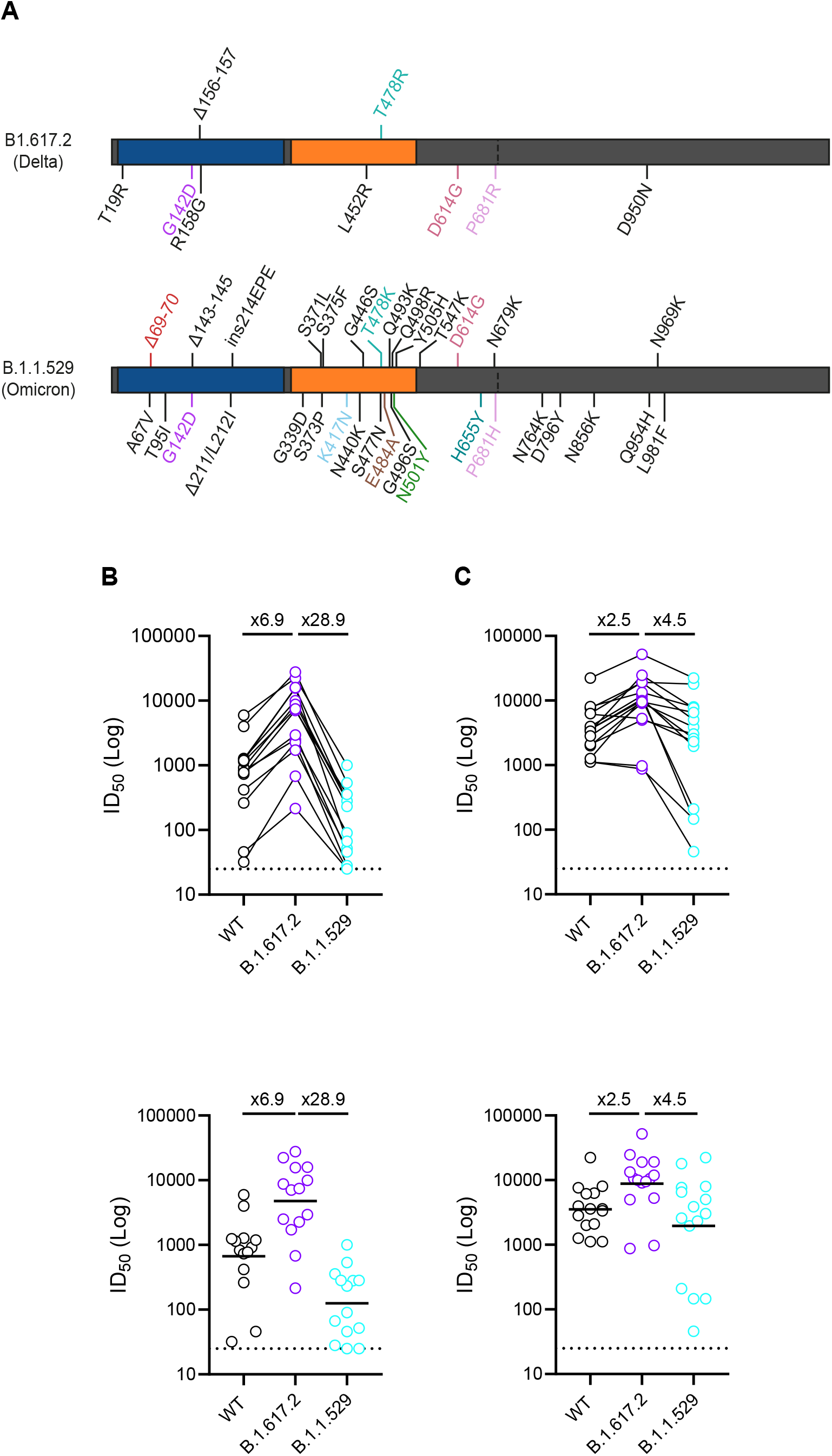
Neutralization of omicron in BTI and delta infected individuals. **A)** Schematic showing mutations in the delta (B.1.617.2) and omicron (B.1.1.529) Spikes. Select sera from **B)** SARS-CoV-2 delta infected (vaccine naïve) individuals (13-22 days POS) and **C)** BTI individuals (12-21 days POS) was tested against WT, delta and omicron VOCs. ID_50_ of neutralization against WT (black) and VOCs delta (purple), and omicron (turquoise) for each participant are linked. Geometric mean titres against WT, delta and omicron VOCs for **D)** SARS-CoV-2 delta infected individuals (13-22 days POS) and **E)** BTI individuals (12-21 days POS). Black line shows the geometric mean titre. Fold decrease in GMT against omicron compared to WT and delta are shown above.

## Discussion

These data demonstrate that whilst 2-doses of COVID-19 vaccine (both BNT162b2 or AZD1222) was not sufficient to provide sterilizing immunity against SARS-CoV-2 infection in these particular individuals, breakthrough infection generated a strong anamnestic response. Although this study cannot provide information on the titre of neutralizing antibody required for protection against infection with the delta VOC, longitudinal sampling revealed that six participants who had undetectable neutralization or ID_50_ ∼25 against delta VOC at the earliest timepoint sampled rapidly developed IgG to Spike and serum neutralizing activity upon infection showing both AZD1222 and BNT162b2 vaccination primed their immune system to respond rapidly upon SARS-CoV-2 infection. 30/42 (71%) of the BTI group were admitted to hospital due to COVID-19 after BTI, the median age was 77 years and only 1/30 (3%) had no comorbidities that would predispose to severe disease. This suggests the group admitted with BTI were at particular risk of severe disease due to advancing age and/or co-morbidities. Indeed, advancing age was the main criterion with which vaccination schedule was based in the UK, meaning that those over 70 years were amongst the first to be offered vaccination in January 2021. As such, vaccine-induced immunity may have waned in this group due to the longer interval between vaccination and exposure, facilitating subsequent BTI (Levin et al., 2021; Mizrahi et al., 2021; Shrotri et al., 2021; Tartof et al., 2021). Indeed, the median time elapsed since last vaccination in the BTI group was 109 days with 24/30 (80%) being vaccinated over 10 weeks prior to symptom onset. Others have described waning of vaccine-induced immunity against delta after 10 weeks, especially in older age groups (Andrews, 2021; Israel et al., 2021). Notably, the unvaccinated group were much younger and a large proportion had no co-morbidities.

When comparing the immune response of the BTI group and the vaccine naïve group, we observe that prior vaccination led to a more potent and broader neutralizing antibody response during the acute phase of infection, including against the highly mutated omicron variant. As we do not have matched sera collected prior to breakthrough infection we cannot comment on the breadth of the nAb response prior to infection. However, in this vaccinated cohort, boosting is occurring with a heterologous Spike which may contribute to the broadening of the serum neutralizing activity. In addition, all individuals in this study received an extended booster regime (8-12 weeks post prime) which has been suggested to generate a broader response than the short (3-4 week) boost regime (Payne et al., 2021; Tauzin, 2021). Further studies examining the antibody response at the monoclonal level is needed to determine if the broader serum activity is due to individual antibodies or a de novo response directed against the delta Spike. Broadening of the neutralizing antibody response has been reported at later timepoints following natural infection (∼6-10 months) (Dupont et al., 2021; Gaebler et al., 2021) and therefore, despite narrow serum neutralization breadth in the vaccine naïve group, convalescent sera collected at later timepoints would be expected to have broader neutralizing activity. The large decrease in neutralization of viral particles pseudotyped with omicron Spike by sera from delta infected individuals highlights the large antigenic distance between the delta and omicron Spike glycoproteins (Dupont et al., 2021; Liu et al., 2021).

Although the omicron VOC is more neutralization resistant, several studies have reported smaller fold-reductions in serum neutralization potency for omicron following 3-doses of COVID-19 vaccination (range 4 - 7 fold) compared to those who had received only 2 vaccine doses (range 20-fold to >40-fold) (Cameroni, 2021; Doria-Rose, 2021; Garcia-Beltran, 2021; Gruell, 2021; Liu, 2021; Schmidt, 2021). Overall, the data presented here suggest that a breakthrough SARS-CoV-2 delta infection is also acting as an effective booster which could provide broad protection against current VOCs, including omicron. As new VOCs arise with new/unique combinations of mutations, our data suggests a broad neutralizing antibody response generated by a combination of vaccination and infection may provide immunity against other/emerging VOCs. This study provides important insights into population immunity and can inform public health measures where SARS-CoV-2 transmission levels remain high.

## Methods

### Ethics

Collection of surplus serum samples was approved by South Central – Hampshire B REC (20/SC/0310). SARS-CoV-2 cases were diagnosed by RT–PCR of respiratory samples at St Thomas’ Hospital, London. Sera were selected on the availability of longitudinal samples and knowledge of timing and type of COVID-19 vaccination.

### COVID-19 severity classification

Disease severity was determined as previously described (Dupont et al., 2021; Seow et al., 2020). Patients diagnosed with COVID-19 were classified as follows: (0) Asymptomatic or no requirement for supplemental oxygen; (1) Requirement for supplemental oxygen (fraction of inspired oxygen (*F*iO2) < 0.4) for at least 12 h; (2) Requirement for supplemental oxygen (*F*iO2 ≥ 0.4) for at least 12 h; (3) Requirement for non-invasive ventilation/continuous positive airway not a candidate for escalation above level one (ward-based) care; (4) Requirement for intubation and mechanical ventilation or supplemental oxygen (*F*iO2 > 0.8) and peripheral oxygen saturations <90% (with no history of type 2 respiratory failure (T2RF)) or <85% (with known T2RF) for at least 12 h; (5) Requirement for ECMO.

### Virus sequencing

Delta variant infection was confirmed using whole genome sequencing as previously described (Dupont et al., 2021) or using MT-PCR (Hale et al., 2021).

### Plasmids

WT, B.1.1.7, B.1.351, B.1.621, B.1.617.2 and B.1.1.529 codon optimized Spike plasmids were obtained from Wendy Barclay (Imperial College London). The final 19 amino acids were removed using an K1255* mutation. B.1.1.7 mutations introduced were ΔH69/V70, ΔY144, N501Y, A570D, D614G, P681H, T716I, S982A, D1118H. B.1.351 mutations introduced were D80A, D215G, Delta242-244, R246I, K417N, E484K, N501Y, D614G, A701V. B.1.617.2 mutations introduced were: T19R, G142D, Δ156-157, R158G, L452R, T478R, D614G, P681R, D950N. B.1.621 mutations introduced were: T95I, Y144T/144insS/Y145N, R346K, E484K, N501Y, D614G, P681H, D950N. B.1.1.529 mutations introduced were: A67V, Δ69-70, T95I, G142D/Δ143-145, Δ211/L212I, ins214EPE, G339D, S371L, S373P, S375F, K417N, N440K, G446S, S477N, T478K, E484A, Q493K, G496S, Q498R, N501Y, Y505H, T547K, D614G, H655Y, N679K, P681H, N764K, D796Y, N856K, Q954H, N969K, L981F.

### Glycoprotein expression and purification

The recombinant wild-type (Wuhan-1 strain) and delta (B.1.617.2) consist of a pre-fusion S ectodomain residues 1–1138 with proline substitutions at amino acid positions 986 and 987, a GGGG substitution at the furin cleavage site (amino acids 682–685) and an N terminal T4 trimerisation domain followed by a Strep-tag II (Brouwer et al., 2020). Spike was expressed in HEK 293 Freestyle cells and purified using StrepTactinXT Superflow high capacity 50% suspension according to the manufacturer’s protocol by gravity flow (IBA Life Sciences).

N protein was obtained from the James lab at LMB, Cambridge. The N protein is a truncated construct of the SARS-CoV-2 N protein comprising residues 48–365 with an N terminal uncleavable hexahistidine tag. N was expressed in *E. Coli* using autoinducing media for 7h at 37 °C and purified using immobilised metal affinity chromatography (IMAC), size exclusion and heparin chromatography.

### Spike IgG titres by ELISA

ELISA was carried out as previously described (Seow et al., 2020). All sera were heat-inactivated at 56°C for 30 mins before use in the in-house ELISA. High-binding ELISA plates (Corning, 3690) were coated with antigen (N or Spike (WT or delta)) at 3 µg/mL (25 µL per well) in PBS overnight at 4°C. Wells were washed with PBS-T (PBS with 0.05% Tween-20) and then blocked with 100 µL 5% milk in PBS-T for 1 hr at room temperature. Wells were emptied and a titration of serum starting at 1:50 and using a 6-fold dilution series in milk was added and incubated for 2 hr at room temperature. Control reagents included CR3009 (2 µg/mL), CR3022 (0.2 µg/mL), negative control plasma (1:25 dilution), positive control plasma (1:50) and blank wells. Wells were washed with PBS-T. Secondary antibody was added and incubated for 1 hr at room temperature. IgM was detected using Goat-anti-human-IgM-HRP (horseradish peroxidase) (1:1,000) (Sigma: A6907) and IgG was detected using Goat-anti-human-Fc-AP (alkaline phosphatase) (1:1,000) (Jackson: 109-055-098). Wells were washed with PBS-T and either AP substrate (Sigma) was added and read at 405 nm (AP) or 1-step TMB (3,3′,5,5′-Tetramethylbenzidine) substrate (Thermo Scientific) was added and quenched with 0.5 M H_2_S0_4_ before reading at 450 nm (HRP). Half-maximal binding (EC_50_) was calculated using GraphPad Prism. Measurements were carried out in duplicate.

### SARS-CoV-2 pseudotyped virus particle preparation

Pseudotyped HIV-1 virus incorporating the SARS-CoV-2 Spike protein (either wild-type, B.1.1.7, B.1.351, B.1 621, B.1.617.2 or B.1.1.529) were prepared as previously described (Dupont et al., 2021). Viral particles were produced in a 10 cm dish seeded the day prior with 5×10^6^ HEK293T/17 cells in 10 ml of complete Dulbecco’s Modified Eagle’s Medium (DMEM-C, 10% FBS and 1% Pen/Strep) containing 10% (vol/vol) foetal bovine serum (FBS), 100 IU/ml penicillin and 100 μg/ml streptomycin. Cells were transfected using 90 μg of PEI-Max (1 mg/mL, Polysciences) with: 15μg of HIV-luciferase plasmid, 10 μg of HIV 8.91 gag/pol plasmid and 5 μg of SARS-CoV-2 spike protein plasmid.(Grehan et al., 2015; Thompson et al., 2020) The supernatant was harvested 72 hours post-transfection. Pseudotyped virus particles was filtered through a 0.45μm filter, and stored at -80°C until required.

### Neutralization assay with SARS-CoV-2 pseudotyped virus

Serial dilutions of serum samples (heat inactivated at 56°C for 30mins) were prepared with DMEM media (25µL) (10% FBS and 1% Pen/Strep) and incubated with pseudotyped virus (25µL) for 1-hour at 37°C in half-area 96-well plates. Next, Hela cells stably expressing the ACE2 receptor were added (10,000 cells/25µL per well) and the plates were left for 72 hours. Infection levels were assessed in lysed cells with the Bright-Glo luciferase kit (Promega), using a Victor™ X3 multilabel reader (Perkin Elmer). Each serum sample was run in duplicate and was measured against the five SARS-CoV-2 variants within the same experiment using the same dilution series.

### Statistical analysis

Analyses were performed using GraphPad Prism v.8.3.1.

## Data Availability

All data produced in the present work are contained in the manuscript

## Acknowledgements

This work was funded by; Fondation Dormeur, Vaduz for funding equipment to KJD, Huo Family Foundation Award to MHM, KJD, MRC Genotype-to-Phenotype UK National Virology Consortium (MR/W005611/1 to MHM, KJD), and Wellcome Trust Investigator Award 106223/Z/14/Z to MHM. CG is supported by the MRC-KCL Doctoral Training Partnership in Biomedical Sciences (MR/N013700/1). This work was supported by the Department of Health via a National Institute for Health Research comprehensive Biomedical Research Centre award to Guy’s and St Thomas’ NHS Foundation Trust in partnership with King’s College London and King’s College Hospital NHS Foundation Trust. This study is part of the EDCTP2 programme supported by the European Union (grant number RIA2020EF-3008 COVAB). The views and opinions of authors expressed herein do not necessarily state or reflect those of EDCTP. This project is supported by a joint initiative between the Botnar Research Centre for Child Health and the European & Developing Countries Clinical Trials Partnership (KJD).

Thank you to Philip Brouwer, Marit van Gils and Rogier Sanders for the Spike protein construct, Leo James and Jakub Luptak for the N protein, Wendy Barclay and Thomas Peacock (Imperial) for providing the Spike plasmids and James Voss and Deli Huang (Scripps) for providing the Hela-ACE2 cells.

