## Supplemental information for "Broad neutralization of SARS-CoV-2 variants, including omicron, following breakthrough infection with delta in COVID-19 vaccinated individuals"

### These authors contributed equally

**Figure S1: A)** Comparison of the IgG and IgM $ED_{50}$ values against recombinant WT and delta Spikes for the AZD1222 and BNT162b2 vaccinated individuals experiencing breakthrough infection. Black horizontal lines show the geometric mean titres. **B)** Longitudinal IgG and IgM $ED_{50}$ against recombinant WT Spike in the BTI group. IgG is show with a circle and IgM is shown with a triangle. Donors with IgM>IgG are shown in blue, donors who do not seroconvert are shown in red, donors with high Spike IgG but no N IgG at <7days POS are shown in orange, and donors with low Spike IgG at <7 days POS that rapidly increases are shown in yellow. **C)** Longitudinal IgG response to N protein. The dotted line represents the cut-off used to determine N seropositivity. **D)** Correlation between $ED_{50}$ against recombinant WT Spike in the BTI group and the optical density (OD) for N IgG binding. The horizontal dotted line represents the lowest dilution used in the neutralization assay. The vertical dotted line represents the cut-off used to determine N seropositivity. Data point are colour coded based on the days POS as indicated in the key. The red box highlights samples that have high IgG binding $ED_{50}$ but low IgG to N.

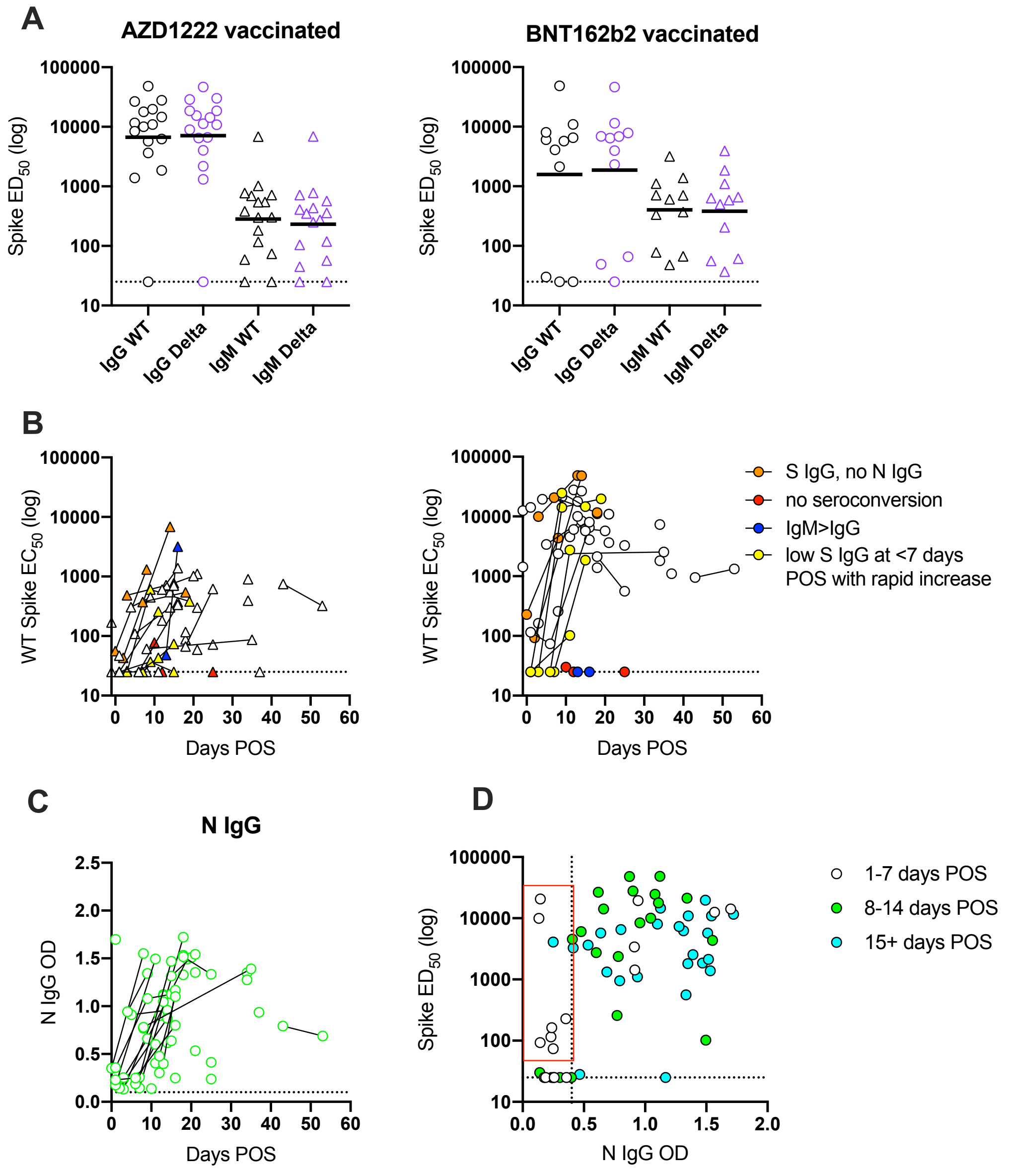

A
AZD1222 vaccinated
BNT162b2 vaccinated
Spike ED50 (log)
IgG WT
IgG Delta
IgM WT
IgM Delta
B
WT Spike EC50 (log)
Days POS
S IgG, no N IgG
no seroconversion
IgM>IgG
low S IgG at <7 days POS with rapid increase
C
N IgG
N IgG OD
Days POS
D
Spike ED50 (log)
N IgG OD
1-7 days POS
8-14 days POS
15+ days POS

**Figure S2: A)** Neutralization potency of sera following breakthrough infection at different days post onset of symptoms (POS) against alpha, beta and mu pseudovirus particles. **B)** $ID_{50}$ of neutralization against WT (black) and VOCs alpha (blue), delta (purple), mu (red) and beta (orange) for sera from AZD1222 vaccinated or BNT162b2 individuals experiencing SARS-CoV-2 delta breakthrough infection. Black line shows the geometric mean titre.

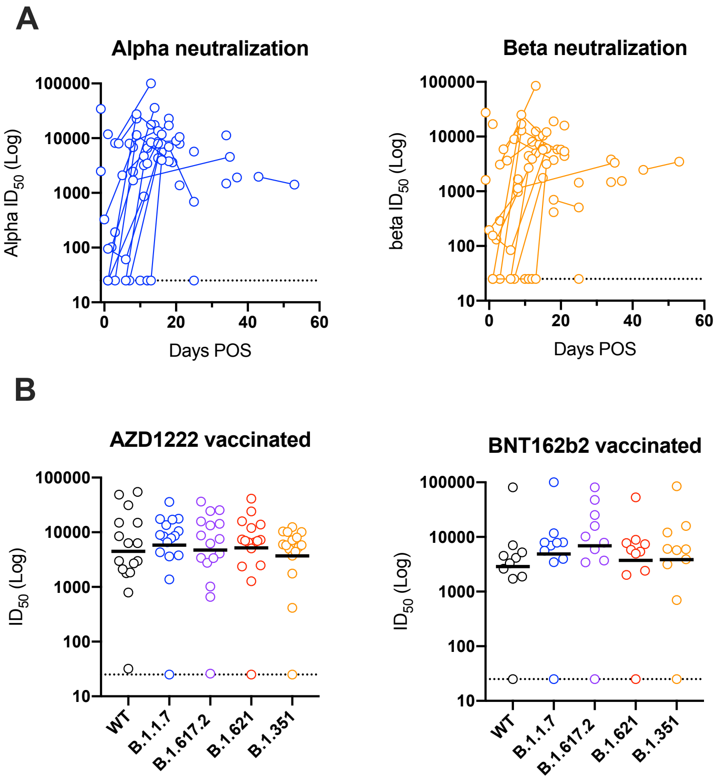
